# Household costs and health-related quality of life of childhood MDR-TB in Western Cape, South Africa

**DOI:** 10.1101/2025.10.14.25338022

**Authors:** Thomas Wilkinson, Anneke C. Hesseling, Graeme Hoddinott, Michaile G. Anthony, H. Simon Schaaf, Edina Sinanovic, James A Seddon

## Abstract

**Background:** Multidrug-resistant (MDR) tuberculosis (TB) in children remains a major public health challenge. Although treatment is provided free of direct charge in many countries, it can impose substantial indirect and non-medical costs on affected households. Evidence on the economic burden of childhood MDR-TB on families, remains limited.

**Methods:** A cross-sectional household survey was conducted in the Western Cape, South Africa, among 45 households with a child <15 years who initiated MDR-TB treatment between 2018 and 2021. Socioeconomic status, costs of accessing care, and health-related quality of life (HRQoL) were assessed and linked to health service utilisation data to estimate household-level costs.

**Results:** The median total cost per household was ZAR 7,443 (US$504) per episode of care (IQR: ZAR 4,119– 13,207), with indirect costs accounting for the largest share of household costs. Twenty-three (51.1%) of the households incurred catastrophic health expenditure, defined as >20% of annual household income. Costs increased with hospital-based care, longer treatment duration, and more frequent caregiver visits. The HRQoL of children was generally high, though not uniformly distributed.

**Conclusions:** Childhood MDR-TB places a substantial financial burden on already vulnerable households. Economic evaluations and care models should incorporate household costs and consider strategies to reduce the indirect burden of treatment on families.

## INTRODUCTION

Multidrug-resistant (MDR) tuberculosis (TB) is defined as disease caused by *Mycobacterium tuberculosis* that is resistant to isoniazid and rifampicin. Globally, an estimated 400,000 individuals develop MDR-TB each year, of whom an estimated 30,000 are children (<15 years)^1^. Treatment of MDR-TB is more complicated than drug-susceptible TB and involves the use of more toxic, and frequently more expensive second-line drugs that typically must be taken over a longer duration. In addition to health system costs, it is critical to understand the costs that are incurred by families for accessing care when a child family member is treated for MDR-TB^1 2^.

While promising research continues to advance the development of shorter MDR-TB treatment regimens and formulations to improve patient experience and outcomes, the available literature on the costs and socioeconomic status (SES) of households with children receiving TB care is sparse. Atkins et al. (2022) conducted a scoping review of studies assessing the socioeconomic impact of TB in children ^3^. While the review identified four qualitative studies focused on the socioeconomic impacts of MDR-TB in children, it did not find any quantitative studies. Shah et al (2023) conducted an extensive SES survey of households where there was a child with drug-susceptible TB (DS-TB) but excluded households with children receiving treatment for MDR-TB given the heterogeneity of treatment pathways and patient experience of DS-TB and MDR-TB^4^. Given that TB disproportionately affects impoverished communities, even minor expenses associated with accessing care can pose substantial barriers for vulnerable households. The absence of data on the financial burden faced by households affected by childhood MDR-TB limits full economic assessment of interventions and treatment modalities, a substantial gap to inform policy. Household surveys can provide granular and localised insights into patient and household experiences of illness across various domains, such as household characteristics, incurred costs, and health-related quality of life (HRQoL) ^5^.

In 2019, the COVID-19 pandemic spread globally and caused substantial disruptions in healthcare access and delivery across various disease areas. Disruptions to TB services impacted on diagnosis, treatment and prevention, with marked increases in disease burden that have persisted for several years post the pandemic in South Africa and in other settings ^6,7^. In addition to better understanding the cost for families of children being treated for MDR-TB, it is also important to understand the impact on these costs of health-system shocks like the COVID-19 pandemic.

## METHODS

### Setting

South Africa has a high burden of TB, with a total TB incidence of 427 per 100,000 population and an estimated 13,000 people with incident MDR-TB in 2023 ^1^. South Africa also faces high rates of child poverty. In 2022, 7.9 million children were living below the food poverty line, and 32% resided in households where no adult was employed ^8^. These households commonly rely on a mix of informal employment and other economic activity, support networks and government grants for income ^9^.

### Study design

A descriptive, cross-sectional household survey was conducted to collect data from households regarding the treatment and impact of childhood MDR-TB. Three survey modules were developed: (a) household SES and employment, (including income and employment related to the COVID-19 pandemic), (b) out-of-pocket expenses for accessing care, and (c) HRQoL. Survey instruments were developed for both paper-based and REDCap digital tools on tablets and were created in isiXhosa, Afrikaans, and English, and participants were matched with researchers fluent in their preferred language.

### Household survey development

The survey was originally adapted from the World Health Organization (WHO) Patient Cost Surveys Handbook (2017)^10^ and a Tuberculosis Control Assistance Program instrument that collected SES information and household expenses of adults with TB (2008) ^11^. Substantial modifications were made, incorporating additional components to align with the study objectives. To collect HRQoL data, permission was gained from the EuroQol group for use and translation of the EQ5D-Y-3L ^12^ instrument into isiXhosa and Afrikaans, and for use of the Toddler and Infant (TANDI) instrument^13^(subsequently renamed the “EQ-TIPS” instrument ^14^).

### Participant recruitment

The study was conducted in Cape Town, Western Cape, South Africa. Clinic records identified 154 households of children who had started routine treatment for MDR-TB when they were below 15 years of age at Tygerberg Hospital between 1 January 2018 and 31 December 2021 and lived in the Cape Town metropolitan area. The study team attempted to contact caregivers by telephone to determine their interest in participating. Upon providing written consent with additional assent from children as applicable, 45 participants (29%) completed the survey in person between March and September 2021. Study staff facilitated participation by collecting participants from their homes or a convenient location. Reasons for not taking part included 11 caregivers (7%) who did not wish to participate, 20 caregivers (13%) who were living out of the area, 9 caregivers (6%) expressed interest but were unable to attend, and 69 caregivers (45%) were uncontactable on the phone numbers available.

### Provincial Health Data Centre

Information on patients’ healthcare resource utilisation (such as the number of appointments and hospitalisations) was extracted from public healthcare records by matching patients’ electronic folder number which serves as a unique identifier. Participants provided their assent and/or consent for researchers to access their electronic health records using this unique identifier through the Provincial Health Data Centre (PHDC). The PHDC, managed by the Western Cape Provincial Government, is an innovative platform integrating diverse per-patient health system data, including hospital admissions, laboratory results, diagnosis, and primary health care details ^15^. A parallel costing analysis in paediatric MDR-TB also used PHDC to synthesise health system costs.^16^

### Analytic approaches

#### Socioeconomic status and income

Household socioeconomic data were characterised by matching reported household assets in the South African Demographic and Household Survey (DHS) 2016, enabling derivation of wealth index scores for each household from the DHS wealth index coefficients. The DHS wealth index has been applied in more than 90 low- and middle-income countries globally and is an established composite indicator that incorporates survey responses on factors such as ownership of assets, dwelling characteristics, water access and sanitation facilities to provide national estimates of relative socioeconomic status ^17^. For this study, household responses were matched to the DHS wealth index coefficients from the general South African population as well as a sub-sample of urban households in the Western Cape, South Africa, offering both national and localised comparisons (supplementary appendix). Information on monthly household income was collected directly via the survey. For those respondents who preferred not to provide household income (n=9), household income was estimated as the median household income of other households in the survey that were in the same SES quintile. Summary statistics of the effect on household income following the COVID-19 pandemic were also recorded.

#### Patient costs

Household survey data were also used to estimate the expected costs of accessing routine care, including visits to hospitals and primary health-care clinics. Direct household costs of accessing care information reported by participants were combined with individual patient treatment profiles, sourced from the PHDC data. Costs are presented in South African Rand (ZAR) 2021 values (the year of collection), and United States Dollars (USD) applying the average exchange rate in 2021 (USD1: ZAR14.78)^18^. For comparison to other country contexts, household costs were adjusted for purchasing power parity and presented in supplementary appendix.

To assess the financial burden of paediatric MDR-TB treatment on households, total household costs related to the child’s MDR-TB care as a proportion of annual household was estimated income. Catastrophic health expenditure (CHE) was estimated per household applying a threshold of 20% of annual household income ^5^.

#### Health-related Quality of Life

Responses from the HRQoL survey module were analysed to generate summary statistics and estimate the health decrements associated with the different stages of the disease. The TANDI^13^ survey was completed by the survey participant on behalf of the child if the child was between 0-<4 years of age (n = 17). The EQ5D-Y ^12^ was completed if the child was between 4-<15 years of age (n=29).

#### Ethics

The study received ethics approval from the Stellenbosch University Health Research Ethics committee no. N20/09/102.

## RESULTS

### Patient Characteristics

Forty-five households and 48 children were included in the survey (**Table 1**). Three households contributed data for two children each. Most households were female-headed (34 families, 76%) and *isiXhosa*-speaking (28 families, 62%), with 15 (33%) reporting Afrikaans and 2 (4%) other languages spoken at home. Household heads were typically a parent of the child 34 (76%), and 30 (67%) had attended but not completed high school. Formal employment was limited, with only 14 (12%) adults employed in the formal sector and 23 (51%) households reporting no employed adult member. Additional health burdens were common: 14 (31%) households included another person with TB, and 8 (18%) reported someone living with HIV in the household; two children (4%) were living with HIV. Among the 48 children, 25 (48%) were <2 years old at the time of TB treatment initiation, and an additional 15 (33%) were 2 to <5 years. The sample was evenly split by sex. Thirty-seven (77%) children had pulmonary TB, while 17 (35%) had extrapulmonary disease.

**Table 1.** Characteristics of survey responses of households with children who had multidrug-resistant tuberculosis (n=45 households)

| Characteristic – household |  | Number (% of households) |
| --- | --- | --- |
| Household Language | isiXhosa | 28 (62) |
|  | Afrikaans | 15 (33) |
|  | Other<sup>†</sup> | 2 (4) |
| Sex of household head | Male | 11 (24) |
|  | Female | 34 (76) |
| Age of household head | 25 to <30 years | 4 (9) |
|  | 30 to <40 years | 17 (38) |
|  | 40 to <50 years | 13 (29) |
|  | 50 to <60 years | 8 (18) |
|  | 60+ years | 3 (7) |
| Relationship of patient to household head | Daughter/Son | 34 (76) |
|  | Grandchild | 6 (13) |
|  | Other<sup>‡</sup> | 5 (11) |
| Highest level of education of household head | Attended but did not complete high school | 30 (67) |
|  | Completed high school | 12 (27) |
|  | Degree or diploma after completing high school | 3 (7) |
| Employment | Households with no adult member employed | 23 (51) |
|  | Proportion of adults in formal employment<sup>#</sup> | 14/113 (12) |
|  | Total unemployment rate of all adults in the household<sup>#</sup> | 77/113 (68) |
| Tuberculosis/HIV | Someone in the household with tuberculosis (in addition to the child) | 14 (31) |
|  | Someone in the household living with HIV | 8 (18) |
| Characteristic – child (n=48) |  |  |
| Age of child | <2 years | 23 (48) |
|  | 2 to <5 years | 16 (33) |
|  | 5 to <15 years | 9 (19) |
| Sex of child | Male | 24 (50) |
|  | Female | 24 (50) |
| HIV | HIV positive | 2 (4) |
| Site of TB | Pulmonary | 37 (77) |
|  | Extrapulmonary | 17 (35) |

<sup>†</sup> Other = English (n=1); French (n = 1)
<sup>‡</sup> Other: Niece/nephew: n = 1; Neighbour/someone I care for: n= 4
<sup>#</sup> Proportion of adults (n=113) in survey households below 65 years of age and who are not in education

### Socioeconomic Status

When household infrastructure indicators for the study sample were compared with national and urban South African averages, a lower proportion of MDR-TB households reported access to electricity compared to the national and urban averages (**Figure 1**). Inadequate housing infrastructure was more common in study households, with 20% residing in informal dwellings compared to 10% of all urban households. Access to piped water inside the dwelling was also lower among study households than the urban average. These findings suggest that households affected by childhood MDR-TB face disproportionate infrastructure disadvantages compared to the broader urban population. COVID-19 had a substantial impact on households, with 24 (53%) reporting that the pandemic had resulted in a major loss of income for the household, and 16 (37%) of caregivers reporting that someone in the house had lost employment due to COVID-19 (supplementary appendix).

**FIGURE 1.**
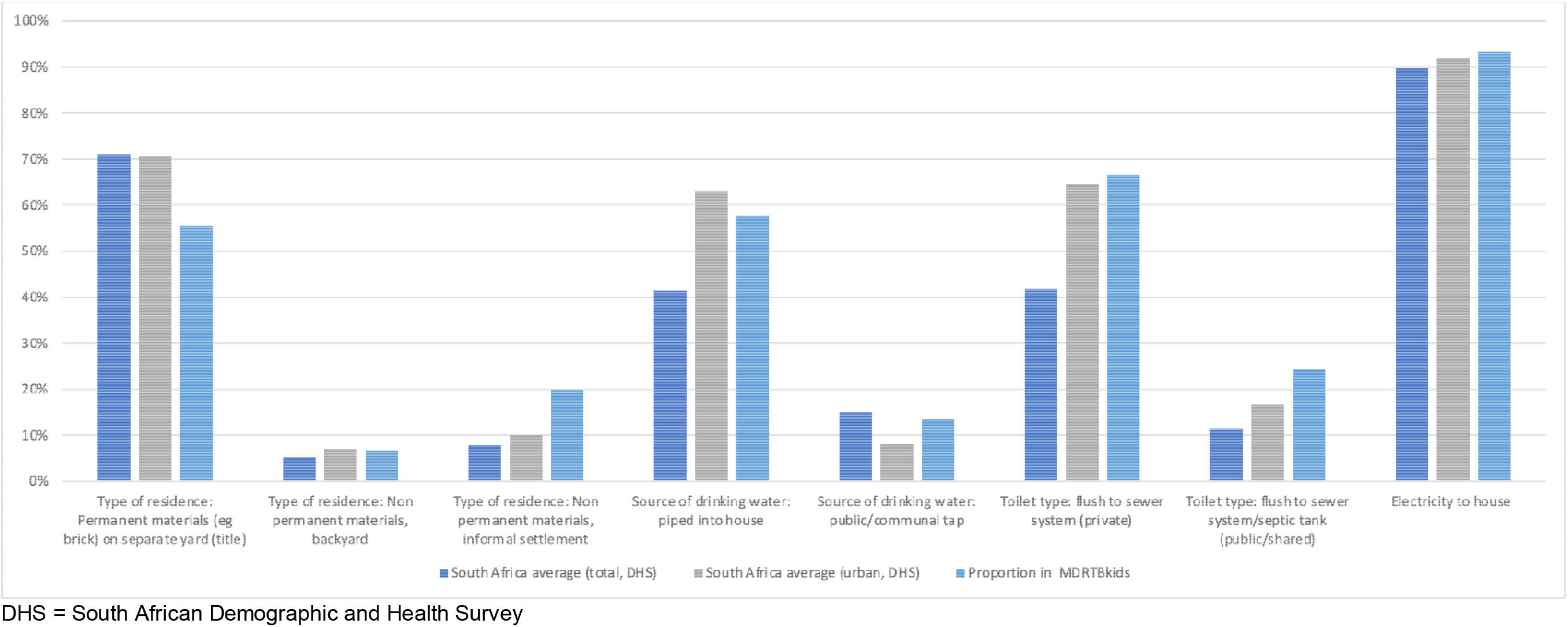
Comparison of household infrastructure in MDRTBkids survey households, all South Arican urban households, and all South African households

### Costs of Accessing Care

For children aged <5 years (n = 39), the median total household cost was ZAR 8,004 (USD 542) (interquartile range [IQR] ZAR 4,310-13,728), including direct medical and non-medical costs of median ZAR 1,949 (USD 132) [IQR: 840–3,430], supplementary costs (which included food purchased while attending health facilities and additional food purchased by the household due to the child’s condition) of median ZAR 2,014 (USD 136) [IQR: 300 – 4,711], and indirect costs related to time and productivity losses of median ZAR 2,959 (USD 200) [IQR: 1,041 – 4,210] (**Table 2**). Among children 5 to <15 years (n = 9), the median total household cost was slightly higher at ZAR 9,130 (USD 618 [IQR: 7,370 -15,416], with indirect costs accounting for the largest share with median ZAR 4,394 (USD 297) [IQR: 2,917–5,357]. The median cost burden as a proportion of household income was 19.4% [IQR: 8%–36%] for households with younger children and 24.9% [IQR: 16–49%] for those with older children. Overall, the combined sample had a median total household cost of ZAR 8,350 (USD 565) [IQR: 4,507–14,133], equivalent to 20.3% [9%–39%] of reported annual household income.

**Table 2.** Median household cost (ZAR) of MDR-TB treatment for a child (interquartile range) by age range, cost type, and proportion of annual household income (n = 48 children)

| Age | Direct costs | Supplementary direct costs | Indirect costs | Total household Costs | Total household costs as proportion of annual household income<sup>†</sup> | Proportion facing catastrophic costs<sup>††</sup> |
| --- | --- | --- | --- | --- | --- | --- |
| 0 to <5 years<br>(n = 42) | 1,949<br>(840 – 3,430) | 2,014<br>(300 – 4,711) | 2,959<br>(1,041 – 4,210) | 8,004<br>(4,310 – 13,728) | 19.4%<br>(8% – 36%) | 47.2% |
| 0 to <2 years<br>(n = 23) | 2,986<br>(1,088 – 3,479) | 1,220<br>(300 – 4,976) | 3,586<br>(2,116 – 5,139) | 9,880<br>(4,870 – 14,766) | 19%<br>(9.2% – 32.0%) | 47.6% |
| 2 to <5 years<br>(n = 19) | 1,050<br>(588 – 2,387) | 3,093<br>(322 – 4,513) | 1,845<br>(976 – 3,469) | 5,540<br>(2,410 – 11,219) | 19.4%<br>(3.8% – 39.0%) | 46.7% |
| 5 to <15 years<br>(n=12) | 2,900<br>(770 – 4,940) | 1,836<br>(1,248 – 6,322) | 4,394<br>(2,917 – 5,357) | 9,130<br>(7,370 – 15,416) | 24.9%<br>(15.5% – 48.7%) | 66.7% |
| Total | 2,016<br>(820 – 3,589) | 1,982<br>(405 – 4,844) | 3,168<br>(1,131 – 4,818) | 8,350<br>(4,507 – 14,133) | 20.3%<br>(8.7% – 39.0%) | 51.1% |

ZAR = South African Rand. MDR-TB = multi-drug tuberculosis. † Mean proportion of annual household income n=45 households.
†† Proportion households where total household costs are greater than 20% of annual household income

Costs were right-skewed, with most households incurring less than ZAR 15,000 (USD 1,015) for the cost of treatment, but a small number reporting much higher expenses. The distribution was approximated by a gamma distribution (shape (▯) = 1.52, scale (▯) = 6,436) (supplementary appendix). More than half (51.1%) of households incurred treatment costs exceeding 20% of their monthly income, a reference threshold for catastrophic health expenditure as defined by the WHO ^5^ (**Figure 2**). Food insecurity, informed by the survey question as “has anyone in the household gone to bed hungry in the previous three months because there was not enough food”, was strongly associated with catastrophic health expenditure.

**FIGURE 2.**
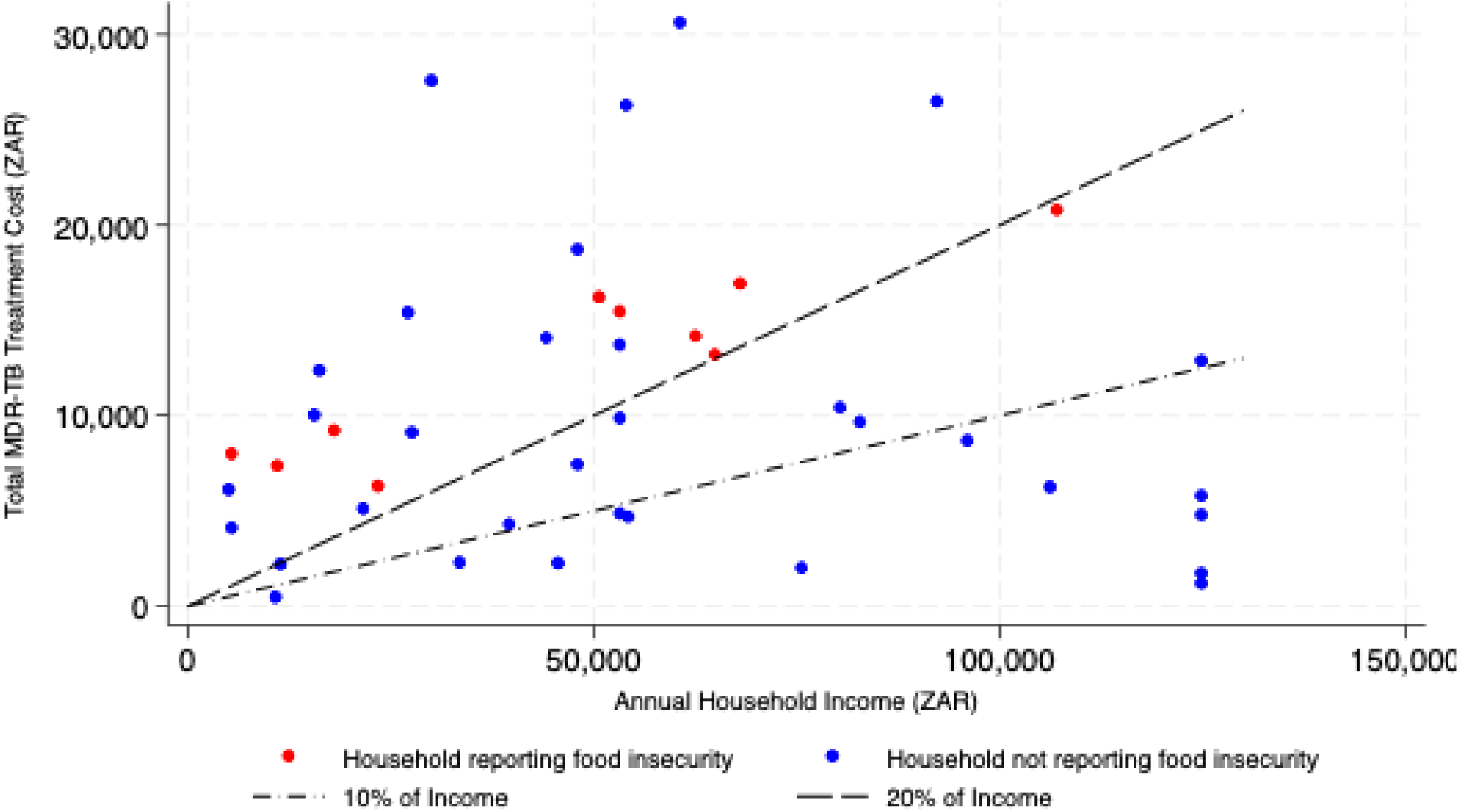
Household costs of accessing multidrug-resistant tuberculosis treatment vs household income with affordability thresholds (n= 45 households)

### Health-related Quality of Life

HRQoL was assessed using the EQ-5D-Y in 29 (63%) children and the TANDI instrument in 17 (37%) children. Most children reported no problems across EQ-5D-Y dimensions, with 27 (93%) having no difficulty doing usual activities and 25 (86%) reporting no problems with mobility or feeling worried or sad **(Table 3)**. However, 7 (24%) reported some or a lot of pain or discomfort. The median EQ-visual analogue scale (VAS) score was 90 (IQR: 80–100), with a mean of 89 (standard deviation [SD]: 13.1). Among the TANDI subsample, problems were most frequently reported in the pain (4 children, 24%) and relationships (2 children, 12%) dimensions. The median VAS score in this group was slightly higher at 95 [IQR: 80–100], with a mean of 87 (SD: 14.4). These results suggest that while most children experienced few limitations in mobility or self-care, pain and emotional well-being were notable concerns for some.

**Table 3.**
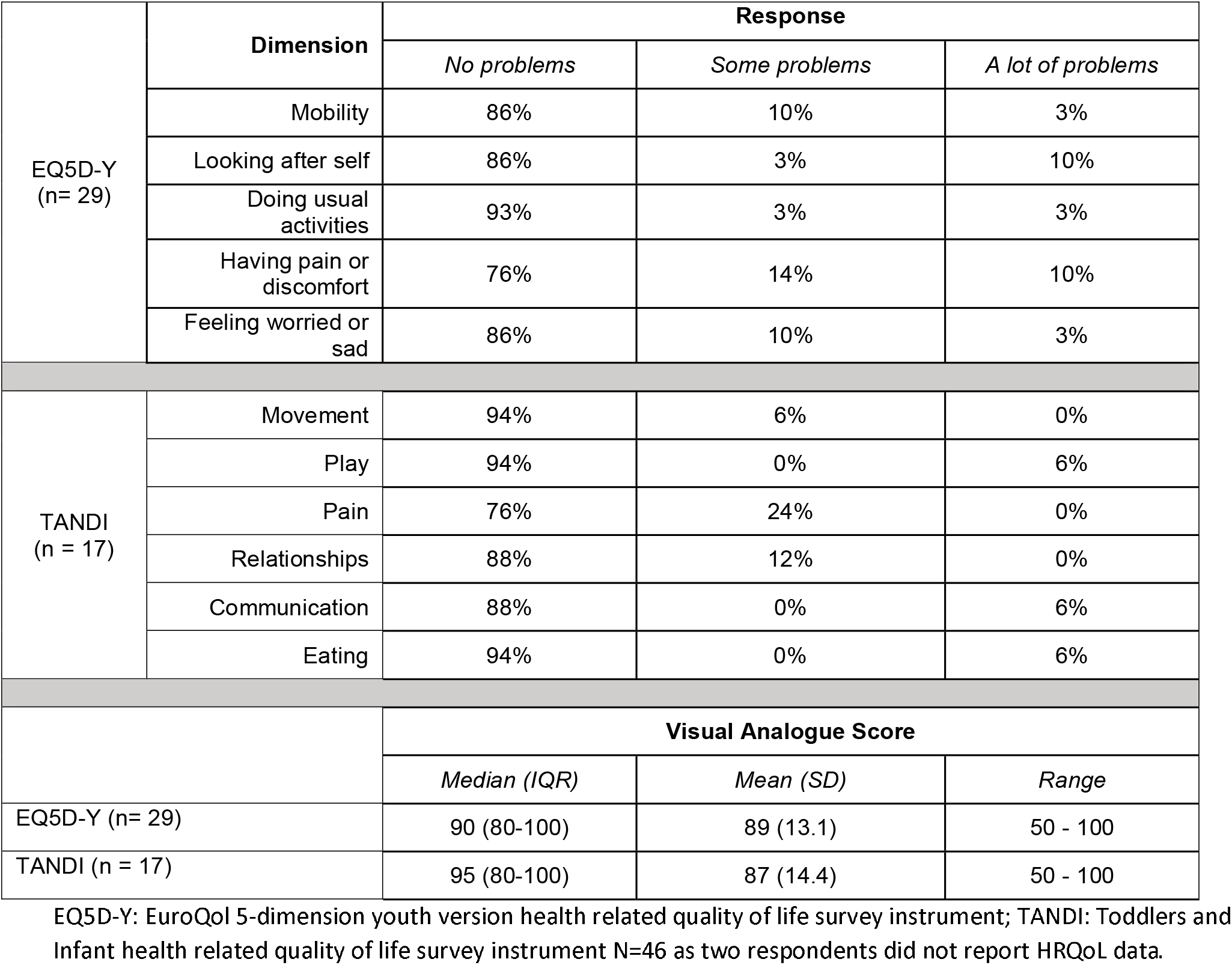
Health-related quality of life of children in the MDRTBkids survey (n = 46)

## DISCUSSION

This study provides one of the first empirical cross-sectional assessments of household socioeconomic status and the cost burden associated with childhood MDR-TB. It captures the real-world experiences of affected households and their interactions with the health system across multiple dimensions. Importantly, it also offers one of the few available descriptions of HRQoL among children with MDR-TB, particularly in the youngest age groups.

A notable strength of this analysis is its integration of primary survey data with a unique routine electronic health information platform in the Western Cape. This approach allows the household “cost per event” (i.e., cost per hospital or primary healthcare clinic visit) to be directly reported by caregivers, while the “number of events” is derived from electronic health records. By separating cost and utilisation data sources, the method reduces recall bias and overcomes common limitations of household costing surveys, where respondents often cannot accurately estimate the frequency of health system interactions across an extended course of treatment. This innovation is particularly relevant for cross-sectional surveys, which otherwise face uncertainty in estimating expected health care utilisation over the full treatment period.

Current WHO guidance on TB patient cost surveys—designed for national programmes—relies primarily on manual data collection from household respondents, with limited reference to the potential use of electronic health records.^5^ While electronic health information systems remain underdeveloped in many high-burden settings, our findings demonstrate that linking routine health records with household-level survey data is both feasible and informative. This provides a new methodological avenue for patient cost studies, with potential to improve precision and reduce data collection burden in contexts where electronic health records are available.

In the study setting, households affected by childhood MDR-TB were disproportionately socioeconomically disadvantaged. Compared to national and urban averages from the 2016 Demographic and Health Survey (DHS), these households had lower rates of formal employment, lower educational attainment, and poorer infrastructure access.^19^ This aligns with global evidence that TB risk is closely linked to structural poverty, but few studies have documented this association specifically in children, particularly in the era of decentralisation for MDR-TB care in provinces such as the Western Cape.

Our findings align with and extend previous research from South Africa. Loveday et al. (2018) reported transport costs of ZAR 93 to ZAR 412 per hospital visit among caregivers of children with MDR-TB, in households earning just ZAR 1,900–2,800 per month ^20^. Ramma et al. (2015) found substantial household costs among adults with rifampicin-resistant TB despite care being nominally free ^21^. Foster et al. (2015) documented mean total costs of ZAR 4,302 per TB episode and noted delays in diagnosis were more common among the poorest patients ^22^. These studies confirm that TB-related costs disproportionately affect poorer households and support the case for stratifying cost data by socioeconomic status—an approach adopted in the present study. These data also indicate the importance of social and economic support for families affected by MDR-TB.

This study also represents one of the earliest applications of the EQ-TIPS, formerly known as TANDI (Toddler and Infant) instrument ^13^ to assess HRQoL among young children (0 to <4 years) with TB. The EQ-5D-Y ^12^ was used in older children (≥4 years), although both tools have limitations in capturing health utility in younger age groups^23^. The findings point to the need for further development and validation of child-specific HRQoL instruments.

Our study had several notable limitations. First, the sample was drawn from an urban referral setting and may not be representative of rural populations or of households with poor outcomes or loss to follow-up—groups likely to face even greater economic strain. Second, uncontactable potential participants may be systematically different (potentially poorer and therefore having less access to a phone) than the sample described. Third, treatment cost estimates were based on observed prices and self-reported utilisation, which may not capture all informal or unrecorded expenditures. Fourth, time costs were valued using the national minimum wage, which may not fully reflect opportunity costs across different employment contexts. Lastly, the HRQoL data were collected for just 46 children without a healthy control group, which limits the ability to determine empirical QoL decrements from this analysis; however, it does provide an important foundation for further research in this population.

The study reinforces the substantial economic and social burden of childhood MDR-TB on already vulnerable households. Even when treatment is provided free of charge, indirect and non-medical costs can lead to catastrophic health expenditure. The study design was unable to establish causal mechanisms to determine whether household poverty increases likelihood of childhood MDR-TB, or whether having a child in the household with MDR-TB increases the likelihood of poverty. The nature of childhood MDR-TB as an expected cause, and symptom, of poverty reflects the complex dynamic between social environment and health. These findings reinforce the need to embed TB services within a broader social protection framework, which may include a range of services including nutrition, transport and financial support, and psychosocial support. In line with the WHO’s End TB Strategy, targeted mechanisms are needed to identify and support vulnerable households, especially given the disproportionate burden of childhood TB among socioeconomically disadvantaged populations.

## Data Availability

All data produced in the present study are available upon reasonable request to the authors

## Acknowledgments

The authors wish to acknowledge the provision of health services utilisation data from the Provincial Health Data Centre, Western Cape Government Department of Health, South Africa.

## Author contribution

TW and JS conceived the survey. TW developed the survey instruments, curated the data (extraction, cleaning, and organization), conducted the analyses, and drafted the manuscript. JS was survey principal investigator. JS and ES provided overall supervision, methodological guidance, and critical revision of manuscript drafts. ACH, HSS, MGA, GH, contributed to survey and study development, design, and analysis, and reviewed and commented on manuscript drafts. All authors approved the final manuscript.

