## Supplementary Appendix 1 for "Household costs and health-related quality of life of childhood MDR-TB in Western Cape, South Africa"

### Supplementary methods

##### 1. Approach to synthesising costs:

Costs experienced by each household were synthesised using survey questions to elicit the cost per event to the household of health service utilisation, and electronic records to extract number of health service events for the household. The cost for each health service event for the household included the return costs of getting to the health facility, money spent while at the facility, and time costs associated with travel and time spent at appointments. For survey participants, electronic records were obtained from the Western Cape’s Provincial Health Data Centre (PHDC), covering the period from 1 January 2018 to 31 December 2022. The advantage of this approach is that the electronic health record is expected to provide an accurate representation of utilisation (i.e. number of PHC visits, outpatient visits, and hospital length of stay), whereas the survey respondents can report their usual costs and time taken for each primary care clinic, outpatient, or hospital visit. Although travel and wait time at facilities is expected to be variable (i.e. some visits incurring longer wait times than others), respondents were requested to report the usual or average time incurred. This method was considered to provide a reasonable reflection of actual household costs from the cross-sectional household survey design. A prospective longitudinal survey involving reported household costs each week or month may have provided more accurate empirical data, however this survey design was not feasible for this analysis.

##### 2. Method for estimating number of health service events for the household:

| **Event type** | **Approach to determining number of event type** |
| --- | --- |
| Primary health care visit | Extract from electronic health record (PHDC) |
| Outpatient visits | Extract from electronic health record  No household event counted if outpatient visit occurred during the child's hospital stay  If multiple outpatient events occurred on a single day, only one was counted under the assumption that the outpatient events could be combined (i.e. a physician visit and an x-ray would be recorded as two outpatient visits in the electronic health record, but if occurring on the same day would be counted as one visit for the purposes of household costs). |
| Hospital admission (general, tertiary, and TB-specialist hospitals) | Extract from electronic health record.  Assumed household event on both day of admission and day of discharge, along with additional visits during the child's hospital stay as follows:  Specialist hospitals: Generally short stay – assume household member/s visit every second day child during the child's hospital stay.  TB specialist: Assume household member/s visit once a week during the child's hospital stay as these were generally of longer duration. |

All events were included if they occurred between the first date of “TB evidence” (a health system event indicative of TB, such as a TB disease code being recorded or drug resistance results) and the TB register date, with an additional allowance of 30 days before and after this period. Where the TB outcome date was not provided in electronic records, length of treatment was estimated based on the median treatment duration for the specific age group in a sample of n= 271 patients in a parallel study utilising electronic records of children with MDR-TB in the Western Cape over the same time period, as reported by Wilkinson et al 2025 (1).

##### 3. Method for estimating cost per event of health service events for the household:

| **Cost type** | **Approach** |
| --- | --- |
| Travel cost | As reported in survey per individual household per event type  Where multiple alternative modes of transport were provided (e.g. bus plus taxi), travel costs were additive. Where travel cost was not reported, median travel cost per event type in the survey sample was applied. |
| Food at event | As reported in survey per individual household per event type.  Where no response supplied, assumed no food expenses. |
| Travel time | As reported in survey per individual household per event type  Where multiple modes of transport provided (for example walk and taxi), the time costs were assumed to be additive.  Where travel time was not reported, median travel time per event type in the survey sample was applied. |
| Time at event | As reported in survey per individual household per event type  Where time at event was not supplied or the survey respondent was unsure, the median travel time per event type in the survey sample was applied.  Time at event for outpatient visits was assumed a fixed value of 60 minutes based on author judgment as this was not collected individually within the survey. |

##### 4. Additional costs as reported by household

*Costs for additional food or supplements.* The survey respondents were asked to state the additional expenditure per month on food or supplements “as a result of the child having TB”, and this amount was multiplied by the length of treatment. It is acknowledged that households may not have consistently spent the reported additional costs for the full treatment period, therefore this cost category is reported separately in the Supplementary results.

*Medical Payments.* One household reported one instance of being required to pay ZAR 75 (USD 4.20) for medicine at a public hospital; there were no other reported costs or required payments (including any additional informal or formal fees, payments or gifts) at public health facilities.

*Cost of non-public sector (private) health provision.* Survey respondents were asked to report both the frequency and cost of visits to non-public facilities such as private doctors, hospitals, pharmacies, traditional healers. This may introduce some recall bias in terms of frequency, however the use of non-public sector providers was relatively small: 7 households reported using a private doctor in general practice (3 visits (n=1 household), 2 visits (n=3 households), 1 visit (n=3 households)); 5 households reported using a community pharmacy (2 visits (n=1), 1 visit (n=3), (one household reported using a community pharmacy 10-12 times during the course of the child’s treatment); 1 household reported visiting a private hospital (where the visit did not result in admission); and one household reported attending a traditional healer on one occasion.

##### 5. Socioeconomic Status (SES) score construction

The household-level socioeconomic status (SES) score used in this analysis was derived using the methodology recommended by the Demographic and Health Surveys (DHS) Program, based on the South African Demographic and Health Survey (SA DHS) 2016. The DHS Program is a longstanding global initiative that has conducted population and health surveys in over 90 countries since 1984. DHS data are widely used to inform analyses of fertility, maternal and child health, nutrition, HIV/AIDS, and related demographic trends.

A standard output of DHS surveys is the household-level wealth index, a composite indicator reflecting cumulative living standards. The wealth index is constructed via principal components analysis (PCA) applied to variables capturing household ownership of durable assets, housing characteristics, and access to water and sanitation facilities. PCA generates a set of factor loadings (coefficients) which are used to weight individual survey items, yielding a standardized continuous score for each household. The DHS wealth index methodology has undergone substantial refinement to enhance cross-population comparability and sensitivity to socioeconomic gradients. All PCA weights and wealth index scores from DHS surveys are made publicly available for secondary use. The full dataset and associated documentation for the SA DHS 2016 are accessible at: <https://dhsprogram.com/methodology/survey/survey-display-390.cfm>.

To construct the SES score for the survey participants, survey items that were identical or comparable between the survey instrument and the SA DHS 2016 were identified (see Table S1.1). The factor loadings from the SA DHS 2016 wealth index PCA were then applied to corresponding survey responses, allowing for the calculation of a standardized SES score for each household in the sample. These scores were then mapped to 1) national South African households (n=11,632) and Western Cape urban households (n= 931) wealth index quintiles defined in the SA DHS 2016, enabling comparison of the relative distribution of survey participants across SES strata, compared to national and Western Cape urban SES wealth scores.

**Table S1: Applicable socioeconomic fields in the MDRTBkids survey used to construct the SES score:**

Socioeconomic questions common to SA DHS 2016 and MDRTBkids survey

Type of residence (dwelling)

Main source of drinking water

Main type of toilet

Electricity to house

Electricity for heating

Electricity for cooking

Asset: Television

Asset: Computer or tablet

Asset: Refrigerator

Asset: Microwave oven

Asset: Oven/stove

Asset: Mobile telephone

##### 6. Health-Related Quality of Life

As the household survey was being completed by the carer of the child with MDR-TB, quality of life was recorded utilising the EQ-5D-Y proxy instrument for patients aged four years and above. Where the child was present and over seven years old, the EQ-5D-Y self-report was also applied. The EQ-5D-Y proxy and self-report instrument has been used extensively to measure health-related quality of life in children (2). The instrument consists of five domains: “mobility”, “looking after myself”, “doing usual activities”; “having pain or discomfort”; “feeling worried, sad, or unhappy”. Each domain consists of three levels, “not/no”, “a bit”, or “very”, for example: “he/she has no pain or discomfort”; he/she has a bit of pain or discomfort”; or he/she has a lot of pain or discomfort”. The EQ-5D-Y self-report aligns to the EQ-5D-Y proxy version where the questions are in the first person (e.g., I have a lot of pain/discomfort”).

The EQ-5D-Y has not been validated in children below 7 years old, therefore the reported HRQoL should be considered indicative only and interpreted with caution (3).

The TANDI (Toddler and Infant Health Related Quality of Life) instrument (4) (subsequently renamed EQ-TIPS) was applied where the child was <4 years. The TANDI instrument is a proxy report (completed by the carer), with six domains: movement, play, pain, relationships, communication, and eating. Each domain has three levels: “no”, “some” or “a lot” – eg within the “eating” domain, options were either “he/she has no problems eating”; “he/she has some problems eating”; or “he/she has a lot of problems eating”.

The EQ-5D-Y and TANDI had a visual analogue scale where the respondent was asked to describe the health of the child today by placing a mark on a scale on a page numbered 0 to 100 where 0 is described as “the worst health that you can imagine” and 100 is described as “the best health that you can imagine”.

As a proprietary instrument, EQ-5D-Y instruments were translated from English into Afrikaans and isiXhosa using the Euro-QoL methodology and validated by the Euro-QoL group. The translations are now available on the Euro-QoL website^[1]^. As the TANDI instrument is non-proprietary for use in research, it was translated to Afrikaans and isiXhosa by the research team with the other modules in the MDRTBkids survey.

### Supplementary results

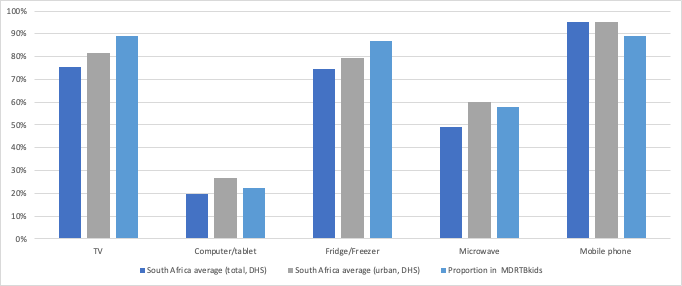

**Figure S1 Proportional ownership of assets: MDRTBkids survey households (n=45), and South African Demographic and Health Survey urban households (n=7,570) and all households (n=11,083).**

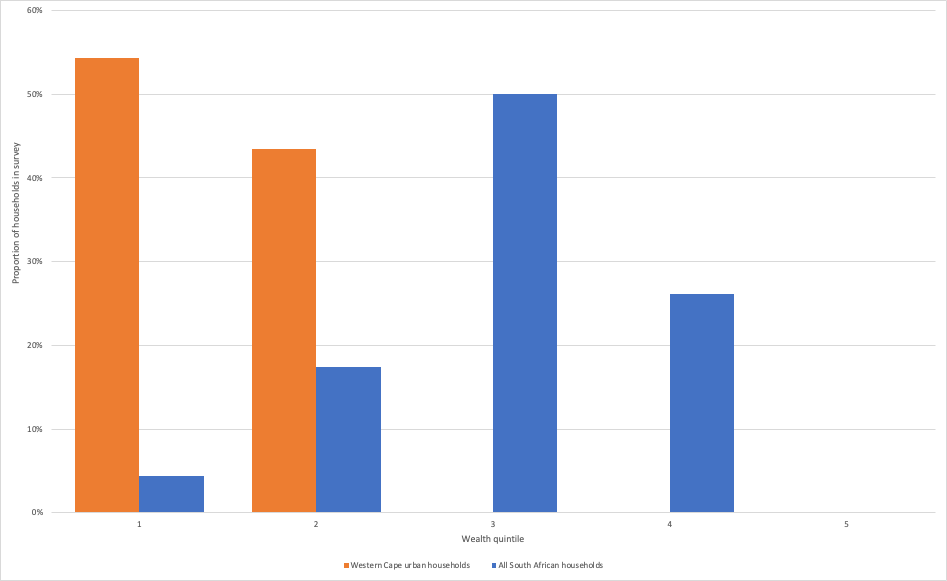

**Figure S2 - Proportion of households in survey (n=45) by wealth quintile**

Figure S2 shows the distribution of survey households by SES index quintile compared to all South African households and specifically urban Western Cape households, based on the South Africa Demographic and Health Survey 2016. The results reveal that, compared to the general South African population, the survey respondents were distributed among the lower four quintiles, with most in quintile three. In comparison with urban Western Cape households, the survey respondents are classified within the lower two quintiles. This difference reflects the generally higher score for urban households than rural ones, especially where household infrastructure and asset ownership are considered, and the relative wealth of the Western Cape province in comparison to other provinces in South Africa.

**Table S2 Median household cost (USD) of MDR-TB treatment for a child (interquartile range) by age range and cost type (n = 48)**

| **Age** | **Direct costs** | **Supplementary direct costs** | **Indirect costs** | **Total household Costs** |
| --- | --- | --- | --- | --- |
| 0 to <5 years (n = 42) | 132 (57 – 232) | 136 (20 – 319) | 200 (70 – 285) | 542 (292 – 929) |
| *0 to <2 years* *(n = 23* | 202 (74 – 235) | 83 (20 – 337) | 243 (143 – 348) | 668 (329 – 999) |
| *2 to <5 years* *(n = 19)* | 71 (40 – 162) | 209 (22 – 305) | 125 (66 – 235) | 375 (163 – 759) |
| 5 to <15 years (n=12) | 196 (52 – 334) | 124 (84 – 428) | 297 (197 – 362) | 618 (499 – 1,043) |
| Total | 136 (55 – 243) | 134 (27 – 328) | 214 (77 – 326) | 565 (305 – 956) |

**Table S3 Median household cost adjusted for purchasing power parity ($I) of MDR-TB treatment for a child (interquartile range) by age range and cost type (n = 48)**

| **Age** | **Direct costs** | **Supplementary direct costs** | **Indirect costs** | **Total household Costs** |
| --- | --- | --- | --- | --- |
| 0 to <5 years (n = 42) | 259 (112 – 456) | 268 (40 – 626) | 394 (138 – 560) | 1,064 (573 – 1,825) |
| *0 to <2 years* *(n = 23* | 397 (145 – 463) | 162 (40 – 662) | 477 (281 – 683) | 1,314 (648 – 1,964) |
| *2 to <5 years* *(n = 19)* | 140 (78 – 317) | 411 (43 – 600) | 245 (130 – 461) | 737 (320 – 1,492) |
| 5 to <15 years (n=12) | 386 (102 – 657) | 244 (166 – 841) | 584 (388 – 712) | 1,214 (980 – 2,050) |
| Total | 268 (109 – 477) | 264 (54 – 644) | 421 (150 – 641) | 1,110 (599 – 1,879) |

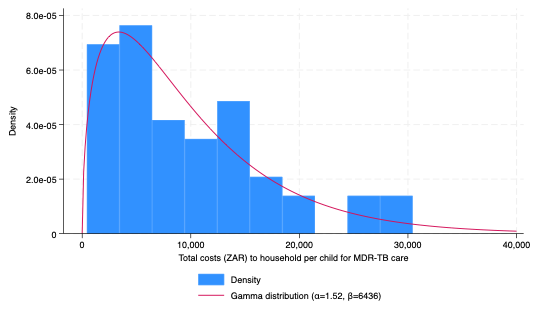

**FIGURE S3: Histogram of total costs (ZAR) to households per episode multidrug-resistant tuberculosis care for a child (n =48) (ZAR)**

**Table S4 Health related quality of life response distribution**

|  | Health state | n | Proportion of sample |
| --- | --- | --- | --- |
| EQ5D-Y  (n= 29) | 11111 | 18 | 62% |
|  | 11121 | 3 | 10% |
|  | 11112 | 2 | 7% |
|  | 12111 | 1 | 3% |
|  | 11113 | 1 | 3% |
|  | 12113 | 1 | 3% |
|  | 12333 | 1 | 3% |
|  | 23112 | 1 | 3% |
|  | 33211 | 1 | 3% |
| TANDI  (n = 17) | 11111 | 11 | 65% |
|  | 111211 | 1 | 6% |
|  | 112111 | 3 | 18% |
|  | 111133 | 2 | 12% |
|  | 232231 | 1 | 6% |

EQ5D-Y: EuroQol 5-dimension youth version health related quality of life survey instrument; TANDI: Toddlers and Infant health related quality of life survey

Table S2.1 above records the scores provided by survey respondents to the dimensions of health-related quality of life in the EQ5D-Y and TANDI instruments respectively. There is a higher proportion in the EQ5D-Y group as the age of the child at the date of the survey determined whether the TANDI (<4 years) or the EQ5D-Y (4 years and above) was used. Where both the proxy and self-complete version of EQ5D-Y was completed, the table above represents the proxy version responses. The Health State reflected in the table above is a score of either 1 (no problems), 2 (some problems), or 3 (a lot of problems), where each number corresponds to the dimensions listed in the table below in the vertical order that the dimensions are listed in the table. For example, a score of 12231 in the EQ5D-Y would indicate no problems with “mobility”, some problems with “looking after self” and “doing usual activities”, a lot of problems with “pain and discomfort” and no problems with “feeling worried or sad”. The survey responses showed that approximately two thirds of children were having no problems in any of the domains, with “pain and discomfort” being the most common dimension with some decrement across both survey instruments.

**Table S5 Health-related quality of life dimensions by instrument**

| EQ5D-Y | TANDI |
| --- | --- |
| Mobility | Movement |
| Looking after self | Play |
| Doing usual activities | Pain |
| Having pain or discomfort | Relationships |
| Feeling worried or sad | Communication |
|  | Eating |

EQ5D-Y: EuroQol 5-dimension youth version health related quality of life survey instrument; TANDI: Toddlers and Infant health related quality of life survey

**Table S6 – impact of COVID-19 on households:**

| Factor | Proportion of sample |
| --- | --- |
| COVID-19 resulted in a major loss of income for the household | 53.4% |
| COVID-19 resulted in a minor or moderate loss of income for the household | 8.9% |
| Someone in the household lost employment due to COVID-19 | 35.6% |

The household survey included brief questions on the impact of COVID-19 and associated lockdowns on household income and employment. More than half the sample (53.4%) noted that COVID-19 had resulted in a major loss of income for the household, and a further 8.9% noted that COVID-19 resulted in a minor or moderate loss of income for the household. In addition, more than a third of households reported that COVID-19 had resulted in at least one person in the household losing employment.

1. <https://euroqol.org/register/obtain-eq-5d/available-versions/>
